# Risk of Type 1 Diabetes in Children is Not Increased after SARS-CoV-2 Infection: A Nationwide Prospective Study in Denmark

**DOI:** 10.1101/2022.12.05.22283089

**Authors:** Rohina Noorzae, Thor Grønborg Junker, Anders Hviid, Jan Wohlfahrt, Sjurdur F. Olsen

## Abstract

**Objective:** It has been hypothesized that SARS-CoV-2 infection in children can increase risk of developing type 1 diabetes.

**Research Design and Methods:** We undertook a prospective analysis based on all children in Denmark where we investigated the association between SARS-CoV-2 infection and subsequent risk of type 1 diabetes, using information from several different national Danish registers. Denmark had one of the highest test-rates per capita in the world during the pandemic.

**Results:** We did not observe a higher risk of a first time diagnosis of type 1 diabetes in children 30 days or more after a positive SARS-CoV-2 test, compared to children with a history of only negative SARS-CoV-2 tests (Hazard ratio 0.85, 95% CI 0.70, 1.04).

**Conclusions:** Our data do not support that SARS-CoV-2 infection is associated with type 1 diabetes, or that type 1 diabetes should be a special focus after a SARS-CoV-2 infection in children.

**Article Highlights:** *Why did we undertake this study?:*

- Studies have shown an association between SARS-CoV-2 infection and subsequent risk of type 1 diabetes, supporting the possibility of a viral etiology in type 1 diabetes and adding to concerns regarding adverse health consequences of COVID-19.

*What is the specific question(s) we wanted to answer?:*

- Is the risk of new onset type 1 diabetes increased among children in the period after SARS-CoV-2 infection?

*What did we find?:*

- We estimated the relative risk of being diagnosed with type 1 diabetes after a positive compared to a negative SARS-CoV-2 test, to 0.85 (95% CI 0.70, 1.04).

*What are the implications of our findings?:*

- Our data do not support an association between SARS-CoV-2 infection and subsequent risk of type 1 diabetes among children.

**Twitter Summary:** A study based on all children in Denmark does not show any association between #SarsCoV2 infection and subsequent risk of #Type1Diabetes among persons < 18 years. #Type1Diabetes should not be a special focus after a #SarsCoV2 infection in children.

---

A number of observational studies have reported an increased risk of diabetes after COVID-19 in persons aged less than 18 years(1–3). This includes two studies on US-based claims databases(1,2) and a study on Norwegian nationwide registers(3). We evaluated the association in Danish nationwide registers.

## Research Design and Methods

Our study was a nationwide, register based cohort study that included all Danish residents aged 0 to 17 years sometime during March 1, 2020 – August 25, 2022 with at least one SARS-CoV-2 test. Inhabitants in Denmark were identified from The Danish Civil Registration System (CPR)(4). SARS-CoV-2 tests (both positive and negative results) were identified in the national COVID-19 surveillance system, which includes all Danish residents with RT-PCR tests for SARS-CoV-2(5).

Type 1 diabetes (DE10) and diabetic ketoacidosis (DE101) diagnoses were identified by ICD-10 codes in The National Patient Register (NPR)(6). The validity of the type 1 diabetes diagnosis among children based on recordings in the Danish NPR has been validated earlier(7). Register information were linked using the unique national ID – the CPR-number – for all Danish citizens. Cohort members were followed from 30 days after the first registered SARS-CoV-2 test and until the end of the study (August 25, 2022) or until the persons either turned 18, died, emigrated from Denmark, designated “missing person” in the CPR register or were censored due to a first diagnosis of type 1 diabetes or diabetic ketoacidosis. Individuals with a registered type 1 diabetes diagnosis or a diabetic ketoacidosis diagnosis prior to study start (March 1, 2020) were excluded.

Hazard ratios (HRs) of being diagnosed with type 1 diabetes comparing follow-up among children with a positive SARS-CoV-2 test to follow-up from children with only negative test results were estimated by Cox regression with current age as underlying time-scale and with adjustment for sex, comorbidity (Charlson’s comorbidity index ≥1, yes/no) at baseline, number of COVID-19 vaccines received (0, 1, ≥2) at baseline, parental history of type 1 diabetes (yes/no) at baseline and current period of year (Jan-Feb, Mar-Apr, May-Jun, Jul-Aug, Sep-Oct, Nov-Dec.). The first 30 days after first positive test were excluded from follow-up. Hazard ratios of being diagnosed with type 1 diabetes with and without simultaneous diabetic ketoacidosis diagnosis according to history of positive test SARS-CoV-2 infection was estimated using a similar approach with a competing risk setup.

## Results

In total 613 cases were diagnosed with type 1 diabetes during 1,593,937 observed person-years among 1,115,716 children aged less than 18 years, corresponding to an incidence rate of 38.5 per 100,000 person-years. Of these, 144 were observed during 419,260 person-years of follow-up among 720,648 SARS-CoV-2 test-positive children. We observed no significant difference in the hazard of being diagnosed with type 1 diabetes in test-positive children compared to children with only negative test results (HR, 0.85, 95% confidence interval CI 0.70, 1.04) (Table 1). We observed similar associations across age, sex, comorbidity, number of vaccine doses, parental history of type 1 diabetes and month of type 1 diabetes diagnosis (Table 1). From 30 days to 6 months since testing positive the HR was 0.88 (102 events, 95% CI 0.70, 1.12) and 0.79 (42 events, 95% CI 0.57, 1.09) more than 6 months after testing positive, compared to children with only negative SARS-CoV-2 tests. The HR of being diagnosed with type 1 diabetes with and without a simultaneous diabetic ketoacidosis diagnosis were 0.61 (17 events, 95% CI 0.35, 1.08) and 0.89 (127 events, 95% CI 0.72, 1.11), respectively. We observed no difference between the estimated HRs in subgroup analyses divided by periods of variant predominance (Table 1).

**Table 1:**
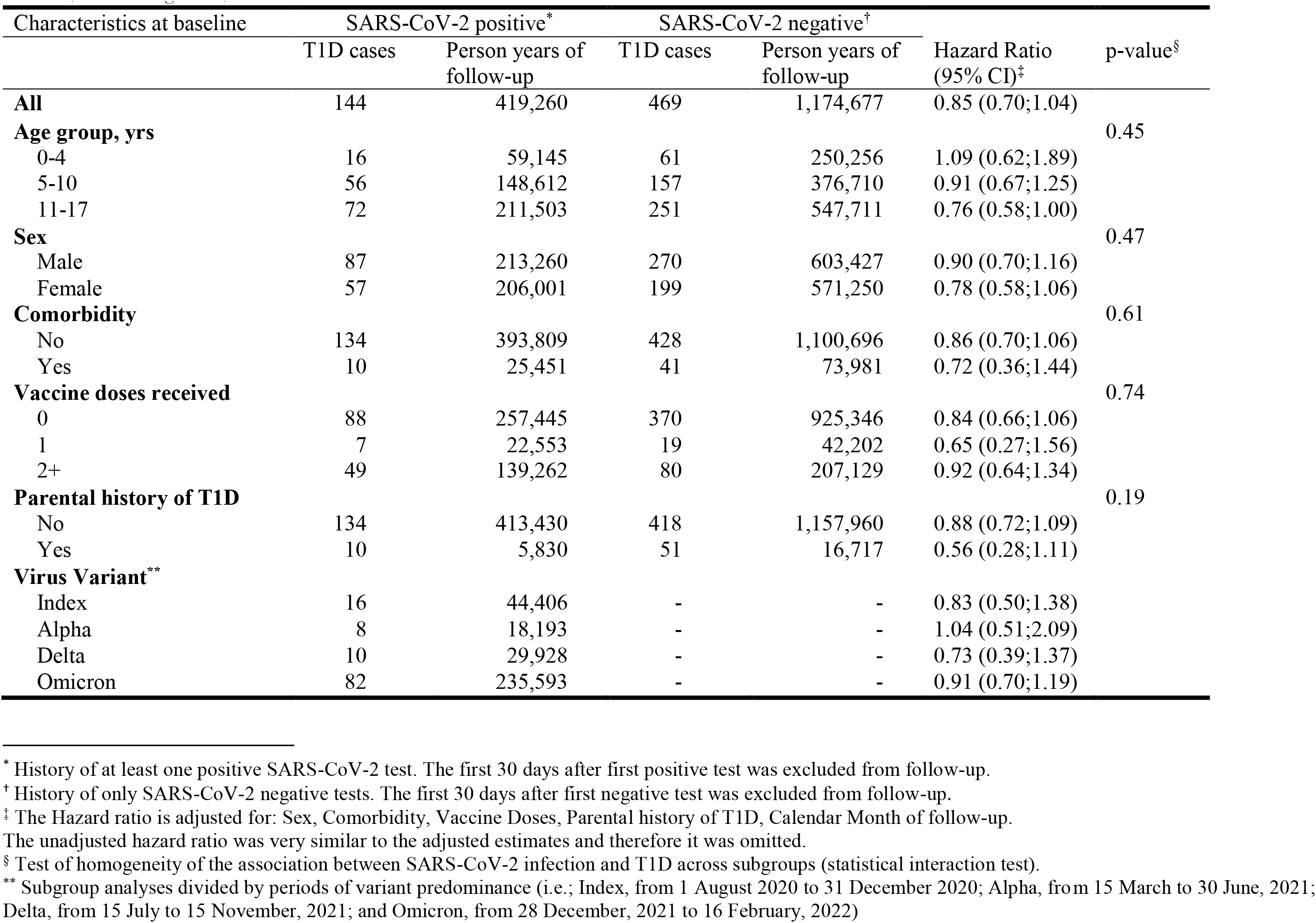
Hazard ratio of Type 1 Diabetes (T1D) by SARS-CoV-2 infection history according to characteristics at baseline in a cohort of 1,115,716 Danish children, March 1, 2020 – August 25, 2022.

| Characteristics at baseline | SARS-CoV-2 positive* |  | SARS-CoV-2 negative† |  | Hazard Ratio<br>(95% CI)‡ | p-value§ |
| --- | --- | --- | --- | --- | --- | --- |
|  | T1D cases | Person years of<br>follow-up | T1D cases | Person years of<br>follow-up |  |  |
| **All** | 144 | 419,260 | 469 | 1,174,677 | 0.85 (0.70;1.04) |  |
| **Age group, yrs** |  |  |  |  |  | 0.45 |
| 0-4 | 16 | 59,145 | 61 | 250,256 | 1.09 (0.62;1.89) |  |
| 5-10 | 56 | 148,612 | 157 | 376,710 | 0.91 (0.67;1.25) |  |
| 11-17 | 72 | 211,503 | 251 | 547,711 | 0.76 (0.58;1.00) |  |
| **Sex** |  |  |  |  |  | 0.47 |
| Male | 87 | 213,260 | 270 | 603,427 | 0.90 (0.70;1.16) |  |
| Female | 57 | 206,001 | 199 | 571,250 | 0.78 (0.58;1.06) |  |
| **Comorbidity** |  |  |  |  |  | 0.61 |
| No | 134 | 393,809 | 428 | 1,100,696 | 0.86 (0.70;1.06) |  |
| Yes | 10 | 25,451 | 41 | 73,981 | 0.72 (0.36;1.44) |  |
| **Vaccine doses received** |  |  |  |  |  | 0.74 |
| 0 | 88 | 257,445 | 370 | 925,346 | 0.84 (0.66;1.06) |  |
| 1 | 7 | 22,553 | 19 | 42,202 | 0.65 (0.27;1.56) |  |
| 2+ | 49 | 139,262 | 80 | 207,129 | 0.92 (0.64;1.34) |  |
| **Parental history of T1D** |  |  |  |  |  | 0.19 |
| No | 134 | 413,430 | 418 | 1,157,960 | 0.88 (0.72;1.09) |  |
| Yes | 10 | 5,830 | 51 | 16,717 | 0.56 (0.28;1.11) |  |
| **Virus Variant\*\*** |  |  |  |  |  |  |
| Index | 16 | 44,406 | - | - | 0.83 (0.50;1.38) |  |
| Alpha | 8 | 18,193 | - | - | 1.04 (0.51;2.09) |  |
| Delta | 10 | 29,928 | - | - | 0.73 (0.39;1.37) |  |
| Omicron | 82 | 235,593 | - | - | 0.91 (0.70;1.19) |  |

\* History of at least one positive SARS-CoV-2 test. The first 30 days after first positive test was excluded from follow-up.
† History of only SARS-CoV-2 negative tests. The first 30 days after first negative test was excluded from follow-up.
‡ The Hazard ratio is adjusted for: Sex, Comorbidity, Vaccine Doses, Parental history of T1D, Calendar Month of follow-up.
The unadjusted hazard ratio was very similar to the adjusted estimates and therefore it was omitted.
§ Test of homogeneity of the association between SARS-CoV-2 infection and T1D across subgroups (statistical interaction test).
\*\* Subgroup analyses divided by periods of variant predominance (i.e.; Index, from 1 August 2020 to 31 December 2020; Alpha, from 15 March to 30 June, 2021; Delta, from 15 July to 15 November, 2021; and Omicron, from 28 December, 2021 to 16 February, 2022)

In a secondary analysis, we looked at the association between COVID-19 related hospitalization and subsequent type 1 diabetes including all individuals 0 to 17 years of age living in Denmark March 1, 2020 – August 25, 2022 (rather than limiting to tested individuals, as in the results reported above). In this extended cohort, we observed a total of 936 cases with type 1 diabetes during 2,817,858 person-years, but we observed no type 1 diabetes cases 30 days or more following a first COVID-19 related hospitalization (939 person-years).

## Conclusions

We did not observe an excess risk of type 1 diabetes following an infection with SARS-CoV-2 in children, such as reported by CDC(1), and subsequently by another study also undertaken in the US(2) and one undertaken in Norway(3). Our results are more in line with a study conducted in Scotland(8). A fifth study was less informative as it did not present estimates of the association for children alone(9).

The matter is important because an association would support a possible viral etiology(10) of T1D and add to already existing worries regarding potential serious adverse long-term consequences of COVID-19 infection.

The CDC study(1) was based two claims databases in the US, viz. IQVIA and Health Verity. From the former, the risk of being diagnosed with diabetes in patients younger than 18 years was estimated to be a factor 2.66 (95% CI 1.98, 3.56) higher when compared to patients of similar age who did not receive a COVID-19 diagnosis during the pandemic; whereas the corresponding relative risk estimated from the Health Verity data was 1.31 (95% CI 1.20, 1.44). As pointed out by others(8), the estimated incidence rates of type 1 diabetes (337 and 351 per 100,000 for IQVIA and Health Verity, respectively) were far higher than could be expected from other sources(8) and approximately 10 times higher than the incidence rate we had estimated for Danish children. One possible explanation(8) might be that a number of prevalent cases had been misclassified as incident cases, leading to overestimated incidence rates. If true, that could potentially invalidate any analysis aiming to identify determinants of diabetes onset based on these data.

The other study from the US(2) was also based on data from a claims database, viz. the TriNetX LLC. For patients 18 years or younger, the risk of being diagnosed with diabetes type 1 within six months of infection with SARS-CoV-2 was a factor 1.83 (95% CI 1.36, 2.44) higher compared with those with non–COVID-19 respiratory infection. Notably, however, the risk was higher already within one month of the SARS-CoV-2 infection by a factor 1.96 (95% CI 1.26, 3.06).

A principal problem common to the two US studies(1,2) was that they both used adjudicated health care claims from primarily commercial health plans. Identifying exposed cases from such databases, and using patients exposed to health problems other than infection with SARS-CoV-2 as reference or comparison groups, can make it difficult to determine what relevant target population the relative risk estimates can be generalized to.

The studies conducted in Norway(3) and Scotland(8), as well as our own Danish study, were based on national health registries for all children and adolescents in the three countries (below 18 years in Norway and Denmark, and below 16 years in Scotland). According to the Norwegian data, the risk of being diagnosed with type 1 diabetes 31 days or more after a SARS-CoV-2 infection, compared to children who had tested negative for SARS-CoV-2 infection, was 1.63 (95% CI 1.08, 2.47). The corresponding relative risk estimated from the Scottish data was 0.79 (95% CI 0.50, 1.27). Thus the Scottish study suggested no association, as did our own estimate based on the Danish data of 0.85 (95% CI 0.70, 1.04). Notably, the confidence interval of our estimate did not overlap with the confidence interval of the Norwegian estimate, as did the much wider confidence interval of the Scottish estimate.

It is not clear why the Norwegian estimate differed from the Scottish and the Danish estimates. Denmark had one of the highest test-rates per capita in the world, and much higher than Norway(11). The proportion of undetected cases of SARS-CoV-2 infection is therefore likely to be higher in the Norwegian than in our study, where 26.3% (419,260PY/1,593,937PY) of the follow-up time was among children who had tested positive for SARS-CoV-2 infection. This fact is also reflected in the substantially higher number of cases identified with an incident type 1 diabetes diagnosis among SARS-CoV-2 infected children in our study, 144, compared to only 28(3) in the Norwegian and 19(8) in the Scottish study.

The confounder distributions might have differed across the three populations, resulting in different risk estimates. Little solid knowledge exists regarding the etiology of type 1 diabetes, apart from the fact that it has a strong genetic component and exhibits substantial familial aggregation(12). Unlike the Norwegian and Scottish study, we were able to adjust for parental type 1 diabetes. This did not impact our estimates.

A strength of our study compared to the four earlier studies(1–3,8) in the field, was that we were able to stratify our data according to periods of SARS-CoV-2 variant predominance. These analyses did not reveal any specific variant patterns.

In conclusion, our data do not support an association between SARS-CoV-2 infection and subsequent risk of type 1 diabetes among persons aged below 18 years, or that type 1 diabetes should be a special focus after a SARS-CoV-2 infection in children.

## Data Availability

Date may be requested by contacting the authors; any request will be subject to The EU General Data Protection Regulation.

## Acknowledgements

No specific funding was obtained for this work.

APH, JW, TGJ, RN and SFO were involved in the conception and design of the study. JW and TGJ were involved in the conduct of analyses. RH, TGJ, APH, JW, and SFO were involved in the interpretation of the results. SFO and RN wrote the first draft of the manuscript, and all authors edited, reviewed, and approved the final version of the manuscript. SFO is the guarantor of this work.

